# Extensions of the SEIR Model for the Analysis of Tailored Social Distancing and Tracing Approaches to Cope with COVID-19

**DOI:** 10.1101/2020.04.24.20078113

**Authors:** Veronika Grimm, Friederike Mengel, Martin Schmidt

## Abstract

In the context of the COVID-19 pandemic, governments worldwide face the challenge of designing tailored measures of epidemic control to provide reliable health protection while allowing societal and economic activity. In this paper, we propose an extension of the epidemiological SEIR model to enable a detailed analysis of commonly discussed tailored measures of epidemic control—among them group-specific protection and the use of tracing apps. We introduce groups into the SEIR model that may differ both in their underlying parameters as well as in their behavioral response to public health interventions. Moreover, we allow for different infectiousness parameters within and across groups, different asymptomatic, hospitalization, and lethality rates, as well as different take-up rates of tracing apps. We then examine predictions from these models for a variety of scenarios. Our results visualize the sharp trade-offs between different goals of epidemic control, namely a low death toll, avoiding overload of the health system, and a short duration of the epidemic. We show that a combination of tailored mechanisms, e.g., the protection of vulnerable groups together with a “trace & isolate” approach, can be effective in preventing a high death toll. Protection of vulnerable groups without further measures requires unrealistically strict isolation. A key insight is that high compliance is critical for the effectiveness of a “trace & isolate” approach. Our model allows to analyze the interplay of group-specific social distancing and tracing also beyond our case study in scenarios with a large number of groups reflecting, e.g., sectoral, regional, or age differentiation and group-specific behavioral responses.

## 1. Introduction

The SARS-CoV-2 pandemic challenges nations worldwide. Since the end of January 2020, many nations around the world have locked down significant parts of their social and economic activities to slow down infection rates. The main goal not to overburden the health system has been achieved with varying degrees of success across countries.

While a lockdown is an effective first reaction to the pandemic, it is obvious that it also comes at significant social and economic costs. Economic forecasts for Germany predict a gross domestic product (GDP) reduction of between 4 and 7 % (approx. 150–260 billion Euros) as compared to 2019 caused by the first month of lockdown only. Beyond the direct economic damage, significant social costs arise in terms of the consequences of unemployment, a sharp increase in the education gap, mental health problems and shorter life expectations due to reduced physical and mental health.

Against this background, one of the most important debates currently is how to overcome lockdowns in a way that maintains a high level of health protection and, in particular, does not overburden the health system. Tailored packages of measures can include (i) protection of risk groups, (ii) suppression of the spread of the virus by legal requirements on protective measures, (iii) suppression of the transmission of the virus by maintaining certain social distancing requirements, (iv) regionally differentiated restarts of social and economic activities, and finally, (v) the improvement of digitalization in the health sector and the use of tracing apps and isolation of potentially infected individuals.

One of the challenges for designing appropriate policies, besides the poor availability of reliable data, is that standard epidemiological approaches do not model key elements of these public health interventions. In this paper, we propose an extension of the epidemiological SEIR model to enable the analysis of commonly discussed measures of epidemic control in more detail. We introduce different groups that may differ both in their underlying parameters as well as in their behavioral response to public health interventions. In particular, we allow the infectiousness parameters to differ within and across groups, which enables the detailed analysis of group-specific measures. Moreover, groups may have different asymptomatic, hospitalization, and lethality rates. Within this framework, we are able to analyze social distancing measures within groups, specific protection of groups with high lethality rates, as well as the detailed effects of “trace & isolate” strategies with different levels of take-up of the tracing apps needed in this context.

In a case study for the German population, we study predictions of our model using parameter estimates from the existing literature in a variety of different scenarios [1–5]. We have to mention at this point that while a lot of experience has been gained over the past year, reliable estimates are not yet available for all parameters. Our model can easily be applied to new and more reliable data sets, as they emerge, as well as richer scenarios with, e.g., a larger number of groups and case studies can be updated as new data become available. On the other hand, our model and results also provide guidance on which parameters are key to be identified reliably in order to analyze the effectiveness of epidemic control approaches in more detail.

Several robust insights emerge from our case study. Our results highlight the sharp trade-off between different goals of epidemic control, namely keeping the death toll as low as possible, limiting the duration of the epidemic, i.e., the time until “enough” immunity exists in the population, and managing intensive care unit (ICU) capacity. We further show that protection of vulnerable groups is effective even if it is applied in a mild form as long as it is implemented in combination with tracing. If tracing is not used together with protection of vulnerable groups, only strict isolation of those groups can effectively prevent a high death toll. Such very strict isolation may be socially undesirable. We also show that compliance is very important for the effectiveness of a “trace & isolate” approach. Tracing is almost completely ineffective unless compliance is high, which in our case study means that at least 70 % of the population install and use tracing apps effectively. Crucially, the “critical” compliance rate can be brought down substantially when vulnerable groups are protected.

Our results suggest that an opening strategy can be effectively supported by the use of apps if they are made mandatory in places where more interaction is allowed. This seems possible at many workplaces and, e.g., in educational institutions, but is probably not effectively applicable in public life in general. Against this background, maintaining group-specific social distancing measures seems inevitable also in the medium run. Our model can be used to study such group-specific measures flexibly for any partition of the population and we re-iterate that any policy conclusion should continually be revised as new and improved data become available. It is also important to note that data protection aspects in connection with tracing apps must be carefully considered, especially as we have seen that high compliance is absolutely essential. Last, extensive digitalization of the health care and the registration system is an indispensable prerequisite for the effective use of a “trace & isolate” approach. The reason is that quarantine of contact persons has to be ordered quick enough to effectively break infection chains.

Epidemiological models such as the classic SIR and SEIR models have been widely applied to model the current crisis (see, e.g., [6–14]) and sometimes have been fitted to the scarce data available [15, 16]. A number of papers have studied public health interventions within the SIR/SEIR models. Most of them focused on social distancing [8, 10, 11]. In general, social distancing is found to be very effective in suppressing the disease in the short run in these models including age-structured and location-specific models [8, 17]. Other interventions have been studied less and, if so, in unstructured models assuming a homogeneous population [18]. Among those, testing and quarantining is seen as a promising tool to control the epidemic [7, 19], though there is widespread awareness of the difficulty of implementing these measures with limited testing capacity and a potentially high asymptomatic rate [3]. Because of this, contact tracing is considered key, but thought to be difficult for COVID-19 and other coronaviruses; see, e.g., [18, 20]. Recent research argues that viral spread is too fast for manual contact tracing in the case of COVID-19 and emphasizes that compliance is critical for the effectiveness of contact-tracing apps [18]. In [21], the authors point out that compliance is also crucial for policies encouraging social distancing. They find that it is essential that a reduction of social activity is implemented robustly on all social groups, especially on those characterized by intense mixing patterns.

One of the contributions of our research is to model the behavioral response, specifically concerning compliance with respect to the use of tracing apps and self-isolation. Our model can furthermore capture sectoral or regional heterogeneity in underlying parameters and infection rates. These properties are also interesting with regard to integration into macroeconomic models, which, e.g., are investigated in [22] or [23].

The remainder of the paper is organized as follows. Section 2 presents the model and Section 3 outlines the scenarios and the parameterization of our case study. Section 4 reports the results of our case study and Section 5 concludes.

## 2. The models

To model the COVID-19 epidemic together with the key aspects of the related political and societal discussion such as the effect of lockdown strategies or tracing apps, we extend the classic SEIR model; see, e.g., [24]. This nonlinear system of ordinary differential equations (ODEs) describes the dynamics of the transitions between four different compartments: susceptible (*S*(*t*)), exposed (*E*(*t*)), infectious (*I*(*t*)), and recovered (*R*(*t*)) individuals. The ODE system is given by

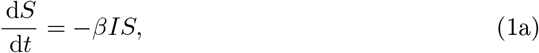

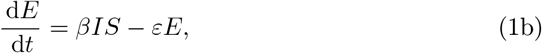

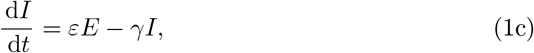

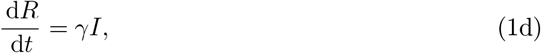

together with given initial conditions

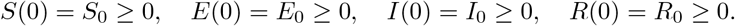

Moreover, it holds *S*(*t*) + *E*(*t*) + *I*(*t*) + *R*(*t*) = 1 for all *t*, i.e., these functions represent fractions of the entire group of considered individuals. Thus, it holds (*S*(*t*) + *E*(*t*) + *I*(*t*) + *R*(*t*))*N* = *N*, where *N* is the total population size. Note that the group of recovered people also contains deaths in this model. The basic reproduction number is *R*_0_ = *β/γ*, which can be thought of as the expected number of cases generated by one case in the model. Table 1 summarizes the notation for the SEIR model.

**Table 1.**
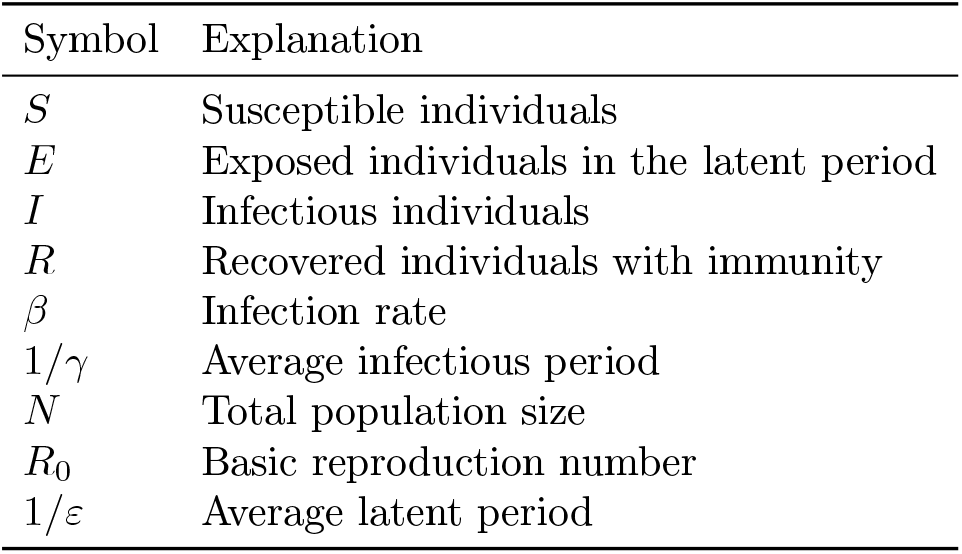
Notation of the SEIR model.

### 2.1. An Extended SEIR Model: SEI^3^RD

The classic SEIR model (1) is now extended to distinguish (i) between different groups such as old and young people, or vulnerable and non-vulnerable individuals, (ii) between different categories of infectiousness (asymptomatic, symptomatic, and severe cases), and (iii) between recovered and dead people. Being able to explicitly distinguish these different groups is important as they can greatly differ in terms of their underlying parameters as well as in terms of their behavioral response to public health interventions.

First, we consider different groups *k* ∈ {1, …, *K*}. The total number of individuals in group *k* is denoted with *N*_*k*_. Groups may differ in multiple characteristics such as age, gender, location, profession, immunity status, etc. The compartments *S, E, I*, and *R* are thus split up into *S*_*k*_, *E*_*k*_, *I*_*k*_, and *R*_*k*_ for all *k*. In our analysis, we also capture specific aspects that are needed to analyze a variety of strategies to slow down an epidemic. We explicitly capture that, across groups, different fractions of infectious individuals show symptoms of the infection. We denote the fraction (of *I*_*k*_) of asymptomatic individuals 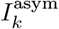 in group *k* by *η*_*k*_ ∈ (0, 1). Thus, 1 − *η*_*k*_ is the fraction of infectious individuals that are symptomatic. Among those symptomatic individuals, a fraction *v*_*k*_ ∈ (0, 1) suffers a severe course of the disease and is assumed to need intensive care unit (ICU) treatment, while a fraction (1 − *v*_*k*_) is symptomatic without the need for intensive care. We denote the group of severely symptomatic infectious by 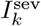 and the symptomatic individuals without a severe course of infection by 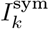. Out of the severely symptomatic infectious individuals in 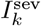, a certain fraction *σ*_*k*_ will not survive. The lethality rate *σ*_*k*_ is group-specific and endogenous, i.e., it depends on the availability of ICU beds and the number of severe cases. We denote the lethality rate given a severely ill patient in group *k* has access to an ICU bed by 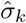. The lethality rate increases endogenously as ICU capacity is exhausted; see Equation (3) below.

In our model, individuals infect other individuals with a certain probability. This probability, modeled via contact rates as in the classic SEIR model, is affected by various aspects, e.g., how likely people meet or whether they show symptoms of the infection. Our model allows for individuals from 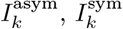, and 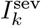 to have different probabilities, depending on their category of infectiousness as well as their group membership. We denote the contact rate between symptomatic individuals of group *k* and individuals of group *j* by 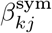, and by 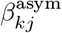 the corresponding contact rate for infectious but asymptomatic individuals. Finally, we also model potentially group-specific average durations of the infection for the symptomatic and severely ill individuals. By contrast, we assume that the average infectious period of asymptomatically infectious individuals is the same across groups.

It is important to note that individuals may start out with an asymptomatic infection but then develop symptoms after a few days or they might initially show only mild symptoms but then develop severe symptoms after several days. We do not explicitly model the transition between these categories, but note that a model extension that would do so is straightforward. In the current model the classes 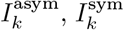, and 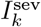 should be interpreted as the expected size of each group at any given point in time. Hence the average infectious period of, for example, symptomatic infectious individuals relates to the time since they became symptomatic.

All notation used in the SEI^3^RD model is summarized in Table 2 and the ODE system of the model is given by

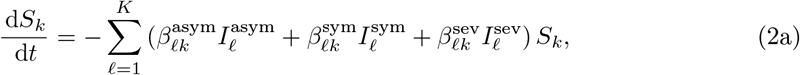

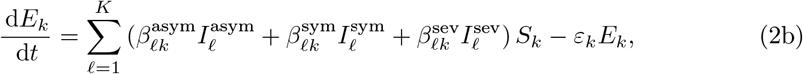

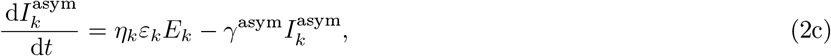

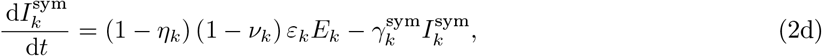

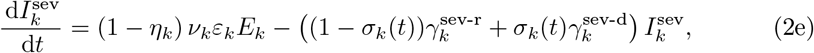

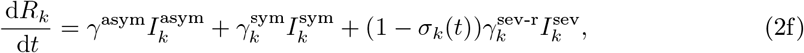

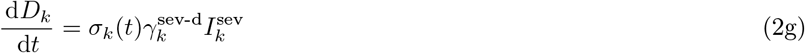

for all *k* = 1, …, *K*.

**Table 2.** Notation used in the SEI^3^RD model.

| Symbol | Explanation |
| --- | --- |
| $K$ | Number of groups |
| $N_k$ | Total number of individuals in group $k$ |
| $S_k$ | Susceptible individuals in group $k$ |
| $E_k$ | Exposed individuals in the latent period in group $k$ |
| $I_k^{\text{asym}}$ | Asymptomatic infectious individuals in group $k$ |
| $I_k^{\text{sym}}$ | Symptomatic infectious individuals in group $k$ |
| $I_k^{\text{sev}}$ | Severely symptomatic infectious individuals in group $k$ |
| $R_k$ | Recovered individuals with immunity in group $k$ |
| $D_k$ | Dead individuals in group $k$ |
| $\eta_k$ | Fraction of asymptomatic infectious individuals |
| $\nu_k$ | Fraction (of $I_k^{\text{sym}}$) of severely symptomatic infectious individuals |
| $\sigma_k$ | Lethality rate conditional on severe infection |
| $\beta_{kj}^{\text{asym}}, \beta_{kj}^{\text{sym}}, \beta_{kj}^{\text{sev}}$ | Group-specific infection rates |
| $1/\gamma^{\text{asym}}$ | Average infectious period of asymptomatic infectious individuals |
| $1/\gamma_k^{\text{sym}}$ | Group-specific average infectious period of symptomatic infectious individuals |
| $1/\gamma^{\text{sev-r}}, 1/\gamma^{\text{sev-d}}$ | Group-specific average infectious period of severely symptomatic infectious individuals who recover (“-r”) or die (“-d”) |

Equation (2a) describes the change in the number of susceptible individuals in group *k* over time. Equation (2b) is the change in the number of exposed individuals in group *k* and *ε*_*k*_ is inversely proportional to the incubation period for individuals in group *k*. Exposed individuals have been infected but are not yet infectious. As specified in Equations (2c)–(2e) we distinguish between asymptomatically (“asym”), symptomatically (“sym”) infectious and severely affected (“sev”) individuals.

A key aspect of the SEI^3^RD model is that the lethality rates *σ*_*k*_(*t*) are endogenous and time-dependent. We assume that, without hospitalization, all individuals with a severe course of the disease die. The number of ICU beds provided by the healthcare system is denoted by *B*. The probability of death and recovery, respectively, depends of the number of free beds in relation to severe cases and also on the selection mechanism that determines who is getting a free bed. We assume that the free beds are rationed proportionally across groups. A different rationing mechanism might be interesting to look at if one is interested in the age structure of the dead or if one wants to analyze the effect of different triage mechanisms that imply preferential treatment, e.g., for young people. Those extensions are easily possible by modifying of Equation (3). Then, *σ*_*k*_ is endogenously determined from the lethality rate 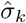 of patients with a severe infection who have access to ICU care as follows,

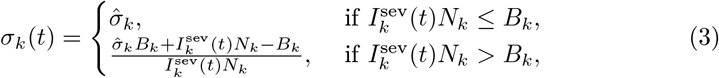

where *B*_*k*_ = (*N*_*k*_*/N*)*B* is the number of ICU beds available to group *k*. The endogenous lethality rate is then finally used in Equations (2f) and (2g) to model the dynamics of the compartments of recovered and dead individuals.

There has been quite some discussion about whether COVID-19 will eventually become seasonal like many other coronaviruses. While our model currently does not allow for seasonality, it can be extended to allow for seasonality or other time heterogeneity in parameters in the same way as the classical SEIR model can be extended to allow for such factors. Both [25] or the literature review in [26] provide good examples of how such seasonality can be modeled.

### 2.2. The SEI^3^Q^3^RD Model: An Extended SEI^3^RD Model to Capture the Effect of Tracing Apps to Break Infection Chains

In the following, we extend the SEI^3^RD model to be able to analyze a policy to reduce the number of infected individuals by the widespread introduction of tracing apps. These apps allow to break infection chains by sending infected individuals to quarantine (*Q*) before they can spread the virus further. Here, we assume that a fraction *ψ*_*k*_ of individuals in group *k* uses such a tracing app effectively. An infection chain can be broken if both, the carrier and the recipient of the virus, use a tracing app “effectively”, meaning that the recipient of the virus can be sent to quarantine before he or she is infectious (i.e., enters one of the groups in compartment *I*). This obviously depends on individual compliance in downloading and using the app but it may also depend on the effectiveness of the health system to contact potentially infected people and send them into quarantine, i.e., on the speed of notification etc. Also note that the fraction of the population that uses tracing apps may vary across groups due to acceptance, ability, or availability of a smart phone. We model the impact of tracing by splitting the compartment *E* consisting of individuals in the incubation period. In particular, we assume that, as a spreader of the virus, only individuals that are aware of their infection (symptomatic or severely infectious) can be recognized by the health authorities and can contribute to break infection chains if they use the app effectively. This applies to a fraction *ψ*_*k*_ of group *k*. All their contacts who also use the app (fraction *ψ*_*Iℓ*_) will be identified. Thus, when individuals from groups *k* and *f* meet, then *ψ*_*k*_*ψ*_*Iℓ*_ of the infected individuals are “traced” and enter 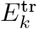, while the remaining infected individuals are “not traced” and enter 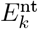. We model the fact that infected individuals who are identified through contact tracing are sent into quarantine by splitting the group of infectious individuals further into *I* and *Q*. All individuals from 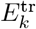 enter compartments *Q* when they become infectious while all individuals from 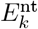 enter compartments *I*. While the individuals in *I* are actively infecting other individuals over time, those in *Q* do not. Individuals from 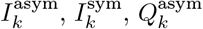, and 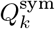 enter the recovered compartment *R*_*k*_ after potentially different durations of illness. Individuals in 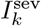 and 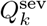 enter either *R*_*k*_ or *D*_*k*_. Again, the lethality rate is endogenous and depends on the availability of ICU beds for individuals in 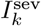 and 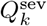; see Equation (5).

The SEI^3^Q^3^RD model looks as follows:

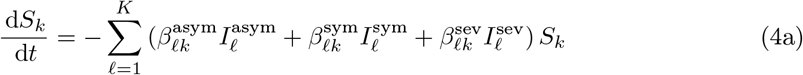

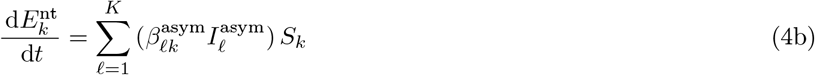

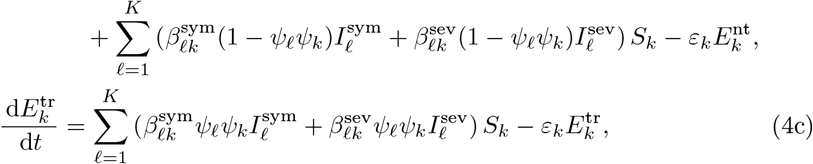

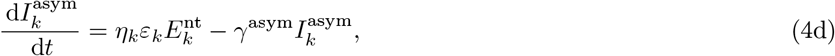

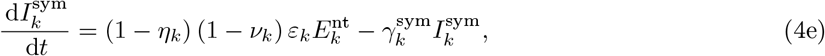

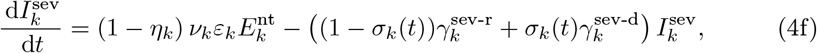

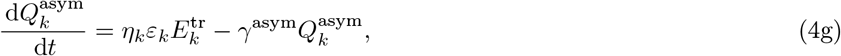

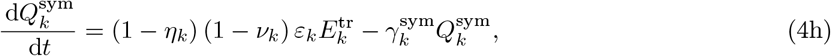

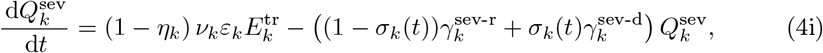

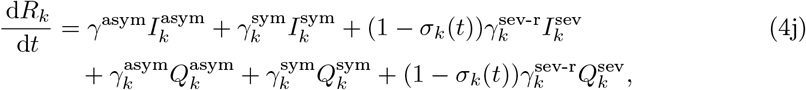

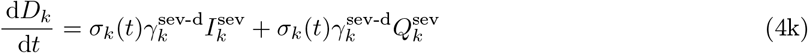

for all *k* = 1, …, *K*.

We again assume that, without hospitalization, all individuals with a severe course of the disease die and we assume again that free beds are rationed proportionally across groups. Then, *σ*_*k*_ is given, in analogy to (3), by

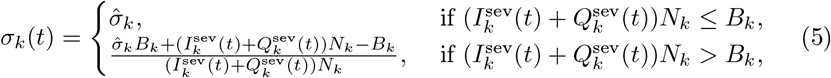

with *B*_*k*_ = (*N*_*k*_*/N*)*B*.

### 2.3. Numerical Solution Approach

The presented SEI^3^RD and SEI^3^Q^3^RD models are nonlinear systems of ordinary differential equations. We solved these systems using an explicit Euler scheme in time (see, e.g., [27]) with an equidistant step size of 10^*−*^_2_. Both the models and the ODE solver are implemented in Python 3.7.4 using the NumPy package. The code and all data to reproduce the results presented in this paper is publicly available at the GitHub repository under https://github.com/m-schmidt-math-opt/covid-19-extended-seir-model.

## 3. Scenarios

We use our models to analyze several widely discussed scenarios and provide evidence on their effectiveness. We provide two benchmarks (laissez-faire and uniform social distancing) and then analyze several variants of more sophisticated epidemic control approaches. The approaches we analyze include (i) specific protection of vulnerable groups, (ii) the use of tracing apps to break infection chains, and combinations of the two. With respect to tracing apps, we consider different take-up (or compliance) rates. All scenarios are also analyzed under different assumptions concerning the asymptomatic rate and the lethality of the virus.

### 3.1. First Scenario: Benchmark

We first consider a benchmark scenario in which no measures are taken. We use estimates of *β, γ*, and *σ* from the literature. In particular, we assume that the values of *β* within and across groups induce a basic reproduction number *R*_0_ between 2.2 and 3. This is the range of typical estimates for COVID-19 [2, 28].

### 3.2. Second Scenario: Uniform Social Distancing

As a second benchmark we consider a standard social distancing scenario in which *β* is uniformly lowered within and across all groups to a value *β*^SD^. We calibrate our model to reach an effective reproductive number of between 0.5 and 1.5 under social distancing in our different scenarios.

### 3.3. Third Scenario: Group-Specific Social Distancing

The third policy that we analyze is to target *β* differently within and across groups. This could, e.g., reflect a policy to largely protect groups that are at risk by implementing measures to lower transmission involving this group. In our parameterization, the vulnerable group is characterized by having a higher conditional probability of a severe course of the disease (modeled using the parameter *ν*) and a higher conditional lethality rate among ICU patients (parameter 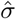). Protection policies for such vulnerable groups are particularly focused on keeping all *β*’s low that involve this group. Interactions within the less vulnerable group can be less restricted. One question we will ask is how much one can allow *β*_LL_, which measures transmission within the low vulnerability group, to increase while at the same time not having to lower the remaining *β*’s too much and still achieve the same effect on overall lethality as under uniform social distancing.

### 3.4. Fourth Scenario: Trace and Isolate

The third widely discussed policy is to reduce the number of infectious individuals *I* by the widespread use of tracing apps that allow to break infection chains. This scenario assumes that a fraction *ψ*_*k*_ of individuals in group *k* uses such tracing apps. The app continuously registers contact persons that an individual comes close to in the context of interactions. Once an individual is tested positively for COVID-19, the individual is free to give the health administration permission to use this data to contact all individuals that have potentially been infected. We assume that all individuals that use the app also agree to the use of their data and that the health office’s procedures are fast enough to actually break the infection chain for all individuals that use the app. This means, in particular, that potentially infected individuals are effectively isolated, i.e., sent to self-quarantine until a test can verify their infection status as positive or negative. Note that the fraction of the population that uses tracing apps may vary across groups due to acceptance, ability, or availability of a smart phone. Note also that the compliance rate can also be interpreted as to reflect the effectiveness of the health system or other aspects that reduce the effectiveness of the app.

As multidimensional policies of infection control we finally also analyze combinations of the measures taken in the third and fourth scenario.

### 3.5. Parameterization

We now describe the parameterization for our case study in more detail. Our case study is modeled to reflect the German situation with a population of about 83 million people. We classify around 17 million people as “high risk”. This approximately corresponds to the share of people aged 65 or older in Germany. Of course some older people are not at high risk, while some younger people with certain health conditions are. The reason to focus on age is (i) that existing data are clear on that age is a very important risk factor and (ii) age, and specifically protection of the elderly, has played an important role in policy debate.

We also emphasize that the aim of the case study is to demonstrate the potential of the modeling approach to provide important insights on the effectiveness of various sophisticated mechanisms of epidemic control. Those mechanisms are important in general to combine effective health protection with social and economic activities. The exact design of the mechanisms will of course depend on the specific situation at hand. A consideration of specific scenarios, with potentially more groups and more accurate parameters is not the focus of this paper, but a possible application of the model. Let us also note that, up to now, we face significant uncertainty concerning key parameters that will drive results and affect policy conclusions.

We now motivate our parameter choices by discussing preliminary evidence on COVID-19 from the very recent literature. If no measures of epidemic control are taken, it is estimated that the basic reproduction number *R*_0_ is somewhere between 2 and 3; see the report [29] by the European Center for Disease Control.

Typical estimates are 3 (reported in [1]), 2.3 (reported in [2]) or 2.26 (reported in [28]). For Germany, the Robert Koch-Institut (RKI) estimated *R*_0_ to be between 2.4 and 3.3 in April [30]. The social distancing measures introduced under lockdown measures across Europe are estimated to induce an effective reproduction number of somewhere between 0.5 and 1. For Germany the RKI estimated *R* = 0.9 with a 95 % confidence interval between 0.8 and 1.1 [30] during the April lockdown. We chose values for our social distancing scenarios that cover this range of estimates and allow also for some milder and some stricter forms of social distancing.

Estimates for the share of asymptomatic infections range from 0.18 on the Princess Diamond cruise-ship, i.e., in a sample of mostly elderly people, see [3], to 0.86 for China [4].

When infected, the incubation period is typically thought to have a median of 5 days—although it can be much faster. The duration of the infection period (after the incubation period and before recovery or death) varies between 7 and 18 days or potentially even longer in severe cases. The severity of COVID-19 differs widely across age groups with less than 1 % of infected under 10 years old being hospitalized but over 15 % aged over 70 needing hospital treatment [5]. Only a subset of the hospitalized need ICU treatment. Based on existing data from Germany [31] we estimate that percentage at around 30 %.

Estimates for conditional lethality vary greatly across existing studies. However, they all highlight a great deal of variation around age. In [32], the authors estimate overall lethality rates of 2.3 % for China (and of over 6 % at that early stage for Italy), ranging from 0.2 % for those under 40 years of age to 14.8 % for those above 80 years of age. [33] study how the age distribution contributes to different lethality rates across countries. In their data overall lethality ranges from 0.7% in Germany to 9.3% in Italy. They find that adjusting for age explains around 66% of the variation across countries with a resulting median lethality rate of 1.9%. The overall lethality rate implied by our parameterization is somewhat lower than these numbers taking into account the lower estimates for Germany and the fact that it is based on *all* symptomatic individuals and not just on officially recorded cases. In a big observational study from hospital patients in Germany during the first wave [31] the authors find that conditional on ventilation mortality ranges from 28% for those aged 18-59 years to 72% for those over 80 years of age. Lethality is substantially lower among those hospitalized who are not ventilated.

For our simulations, we split the population in two groups. One group is assumed to be composed out of people having a high risk of a severe course of the infection, the other group having a lower risk. We systematically vary the parameters *β, η*, 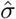, and *ψ* to gain insights into the effect of social distancing, the protection of the vulnerable group, the impact of the asymptomatic rate, and the effect of the use of a tracing app to break infection chains. Apart from this, all scenarios are based on the parameterization given in Table 3, which summarizes our fixed parameters.

**Table 3.** Parameter values.

| Description | Parameter | Low Risk | High Risk |
| --- | --- | --- | --- |
| Average incubation time | $1/\varepsilon$ | 5 | 5 |
| Infection duration, asymptomatic | $1/\gamma^{\text{asym}}$ | 10 | 10 |
| Infection duration, symptomatic | $1/\gamma^{\text{sym}}$ | 12.5 | 12.5 |
| Infection duration, severe & recovered | $1/\gamma^{\text{sev-r}}$ | 20 | 20 |
| Infection duration, severe & dead | $1/\gamma^{\text{sev-d}}$ | 14 | 14 |
| Share of ICU patients among<br>the symptomatic | $\nu$ | 0.0075 | 0.045 |
| ICU beds | $B$ | 24 000 | |
| Total population size | $N$ | 66 119 010 | 17 041 997 |

Table 4 shows the parameters that change across the various scenarios that we simulate. Parameter values are chosen to be in line with estimates of the basic reproduction number and to cover interesting cases. We will further motivate specific parameter choices in the course of the presentation of our results. In the benchmark (B) and the uniform social distancing (USD) scenarios, we have chosen uniform *β* parameters across groups. For the group-specific social distancing (GSD) scenarios, we split the population in a vulnerable and a non-vulnerable group and choose different *β* parameters for interactions that involve the vulnerable group and those that do not. We moreover vary the lethality rates and the asymptomatic rates to gain some insights into the sensitivity of the results.

**Table 4.**
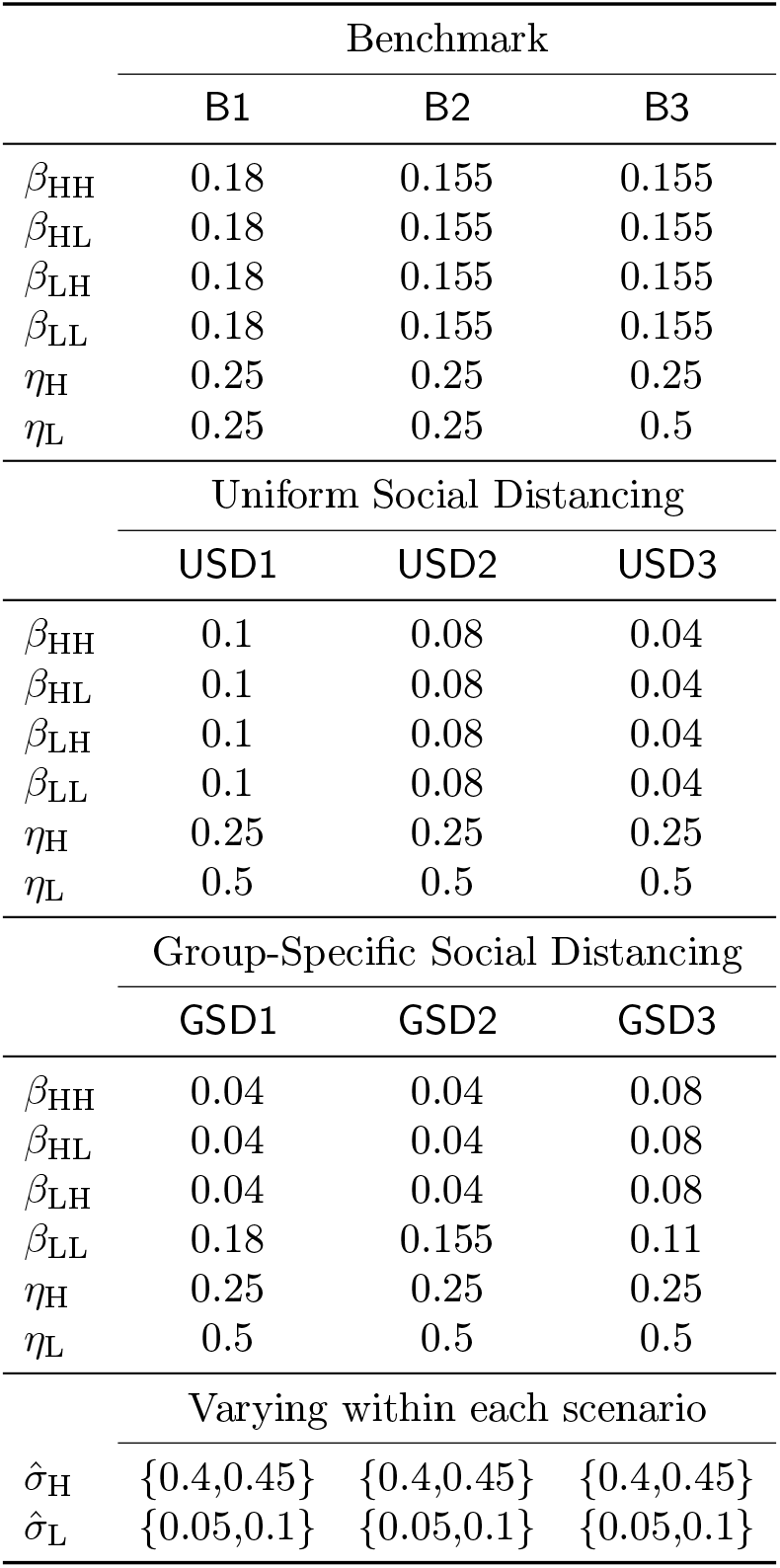
Parameters that vary across the main scenarios discussed below.

## 4. Results

This section contains the results and discussion of our simulation outcomes. In order to first draw the bigger picture, we present two selected scenarios that illustrate extreme cases, the scenarios B2 and USD2 (see Table 4). Scenario B2 represents a situation without any measures of epidemic control and a reproduction rate of *R* ≈ 2.6. Scenario USD2 assumes thorough social distancing leading to an effective reproduction rate *R* ≈ 1. Note that since asymptomatically, symptomatically, and severely infectious individuals can have different recovery rates in our model, *R* changes over time as the relative frequency of these groups in the population changes.

This is why the values of *R* we indicate are only approximations. The course of the epidemic in the two scenarios is presented in Figure 1 with days since the start of the epidemic on the *x*-axis and number of individuals on the *y*-axis. Note that for both scenarios we show group-specific outcomes for a group with high vulnerability and a group with low vulnerability (L). Here and in what follows, we define the duration of the epidemic as the number of days until the number of severe cases is below 5 % of ICU capacity and where at the same time the share of the population in the susceptible category is below 1*/R*_0_.

**Figure 1.**
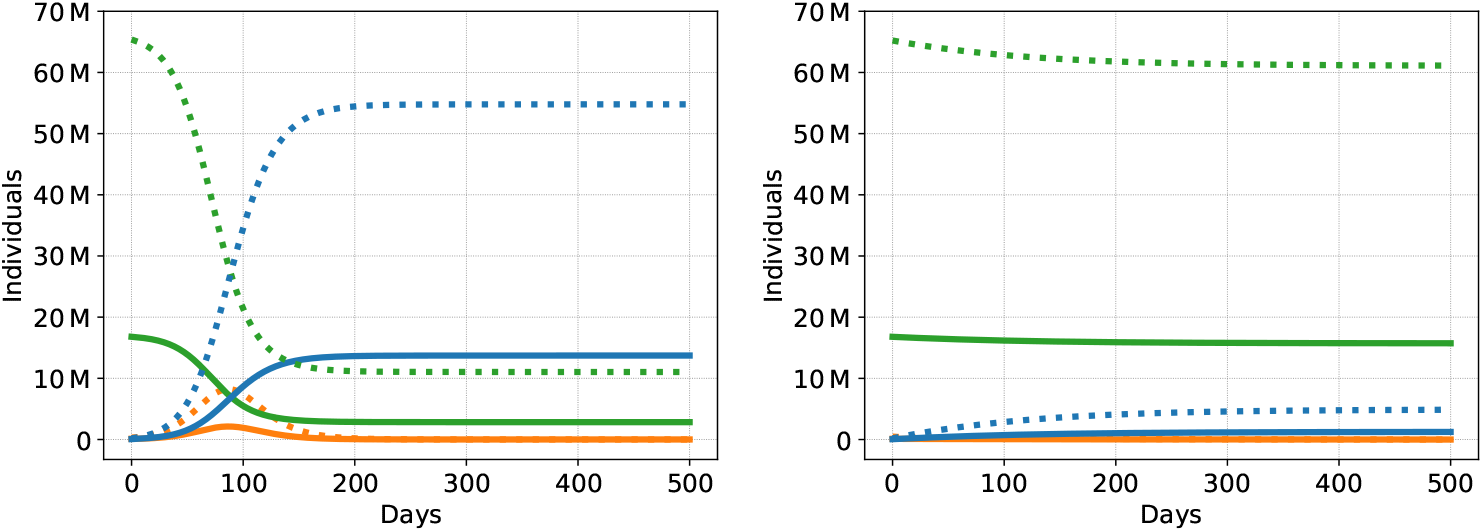
Dynamics of the COVID-19 epidemic for the B2 (left) and the USD2 (right) scenario: Susceptible individuals in green, recovered in blue, and aggregated asymptomatic, symptomatic, and severely infectious cases in orange. The group with low vulnerability has dotted curves, the group with high vulnerability is plotted solidly.

Figure 1 (left) illustrates that without any intervention, the epidemic is quickly over but claims many lives. In particular, due to the rapid exponential spread of the epidemic in scenario B2, about 80 % of the population has gone through the infection after only 247 days. At this point, only about 20 % of the population remain susceptible. Due to the fact that the health system’s ICU capacity is exceeded for 140 days during the epidemic, more than 500 000 individuals die. Due to scaling, this is not visualized in Figure 1 but we discuss the details in Section 4.1.

By contrast, under social distancing, see Figure 1 (right), the epidemic lasts far longer than 500 days. After the first 500 days, 92 % of the population is still susceptible. On the positive side, the health system’s capacity is always sufficient and only 26 000 individuals die. On the negative side, the epidemic has not been overcome: The situation faced by society after 500 days is similar to the one on day zero.

These figures clearly illustrate that both options are no realistic alternatives for a society to deal with the epidemic in the medium run. Scenario B2 implies an unbearable situation for society, while USD2 does not solve but preserve the problem, with the solution still pending and the economy shut down with all the associated problems. In the following, we thus discuss these two and other benchmark scenarios (Section 4.1) and social distancing scenarios (Section 4.2). We then illustrate what our model can contribute to the understanding of the effectiveness of more sophisticated and tailored approaches of epidemic control. In particular, we focus on group-specific social distancing (Section 4.3) and the effect of tracing apps (Section 4.4), as well as combinations of the two.

### 4.1. Benchmark

We first analyze three benchmark scenarios in which we assume that no epidemic control takes place. For details and exact parameterizations see Table 4. Scenario B1 is characterized by a high reproduction number (on average *R* ≈ 3) and a low asymptomatic rate of 25 % for both groups. In B2, *R* ≈ 2.6 and otherwise the parameters are identical to B1. In B3, *R* ≈ 2.2 and the L group has a higher asymptomatic rate of 50 %. Each model is run using varying assumptions on lethality rates; see Table 4.

For each model, we report in Table 5 the death toll within the first 500 days of the epidemic, the number of days in which ICU capacity is exceeded, the duration of the epidemic, and the fraction of susceptible individuals after 500 days.

**Table 5.** Results for the Benchmark Scenarios

|  | B1 | B2 | B3 |
| --- | --- | --- | --- |
| Deaths in first 500 days | 537–627 K | 445–527 K | 238–329 K |
| ICU capacity exceeded on | 125–132 days | 138–140 days | 144–150 days |
| Duration | 219–239 days | 241–247 days | 273–281 days |
| Susceptible after 500 days | 10–12 % | 19–21 % | 21–23 % |

In line with other studies [9, 10, 34], our results suggest that the epidemic will claim a large number of lives if left unchecked. In our simulations, ICU capacity would be exceeded for between three and five months and the death toll would be anywhere between 240 000 to over half a million lives. Because of these and similar results we have seen social distancing implemented all over Europe in the last few months. Against this background, the next section addresses various social distancing scenarios.

### 4.2. Uniform Social Distancing

As a second benchmark we consider three uniform social distancing scenarios. Uniform social distancing means that all interactions in the population are reduced to the same extent. Among our three scenarios, USD1 represents a case of mild social distancing with an average effective reproduction number of *R* ≈ 1.5. All parameters but the contact rates *β* are identical to B3. In this scenario, the epidemic is slowed down but the number of cases is still increasing during the first 500 days. Scenario USD2 has *R* ≈ 1, i.e., also here the disease is progressing, but more slowly. Scenario USD3 suppresses the disease with *R* ≈ 0.6, which is at the lower end of the estimates of the effective reproductive number during the recent/current lockdowns in Europe.

Under mild social distancing (USD1) the epidemic progresses slowly. ICU capacity is slightly exceeded over a period of at most 11 days, but there is still a death toll of over 120 000 in the first 500 days. After 500 days, a large share of the population is still susceptible and, moreover, it is unknown, given current evidence, whether those who passed through the illness early on are still immune after 500 days. In USD2 and USD3, social distancing is effective to suppress the epidemic with the effect that the number of deaths in the first 500 days is comparatively low. However, in these scenarios, the vast majority of the population (up to 99 %) is still susceptible after 500 days. Hence, especially in USD2 and USD3, the death toll will further increase after 500 days until a vaccine is widely available. Note also that all our uniform social distancing scenarios are based on the benchmark scenario B3, which assumes a higher asymptomatic rate among the group with low vulnerability. In the case of a lower asymptomatic rate (comparison to B2), there are more deaths in the first 500 days. Other than that the results are very similar. Suppression is unlikely to be an effective long term strategy and, in the absence of a widely available vaccine, alternatives need to be considered. This is what we focus on in the next two sections.

### 4.3. Group-Specific Social Distancing

Our group-structured model allows us to study non-uniform social distancing measures, which are designed to induce different within-group and across-group contact rates. This could allow to explicitly protect vulnerable groups (as shown in our case study) or to design tailored social distancing measures that allow particularly desirable or important economic and societal activities while suppressing others. This means, groups could be defined by sector, age, region, or otherwise.

In this part of our case study we focus on one particular split, which has received much attention in recent policy discussions in which the idea is to protect vulnerable groups. We hence split the population in the 20 % most vulnerable and the rest of the population. The 20 % most vulnerable individuals are those most likely to have a severe course of the disease. It has been shown that mostly age, but also gender and certain health conditions, are strong predictors of the severity of COVID-19. Of course, it is also possible to split the population in other ways and, e.g., to protect the most vulnerable 25 or 30 %.

The scenarios we discuss here build on our benchmark scenarios B2 and B3. We then either reduce only contact rates for interactions that affect individuals with high vulnerability (H) or reduce all contact rates but to a different extent. The first approach corresponds to a strategy to resume life as usual except for the vulnerable, who are then severely protected. The second approach corresponds to the establishment of general protection measures also in daily life, with particular protective measures applied to vulnerable individuals. GSD1 builds on benchmark B2, but then drastically reduces all interactions involving highly vulnerable individuals, i.e., contact rates to *β*_HH_ = *β*_LH_ = *β*_HL_ = 0.04. This roughly corresponds to an effective reproduction number of *R* ≈ 0.6 generated from these interactions only. GSD2 also restricts all interactions involving the highly vulnerable population to *β* = ≈ 0.04 but corresponds to B3 in all other parameters, i.e., in particular assumes a lower contact rate *β*_LL_ from interactions among the less vulnerable. GSD3 is a scenario where both groups engage in social distancing but to a different extent, with LL-interactions restricted to *β*_LL_ = 0.11 and all other interactions restricted to *β*_HH_ = *β*_LH_ = *β*_HL_ = 0.08. Otherwise, the parameters in GSD3 are identical to GSD2 and B3. Table 7 reports the results for these three scenarios.

**Table 6.**
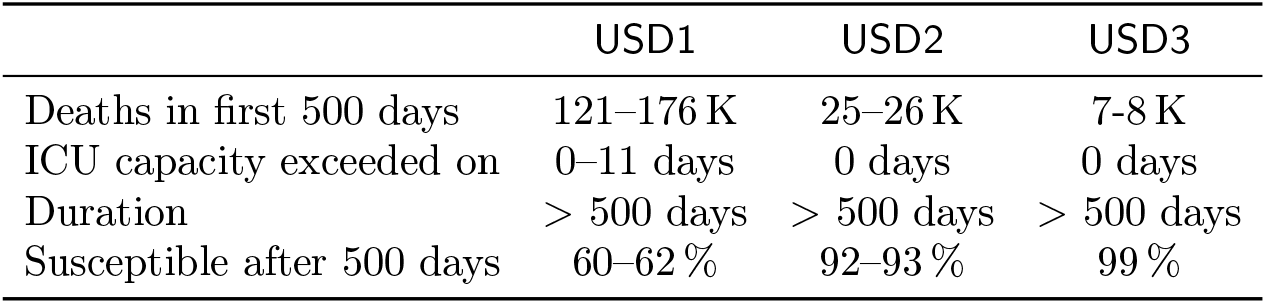
Results for the Uniform Social Distancing Scenarios

**Table 7.** Results for the Group-Specific Social Distancing Scenarios

|  | GSD1 | GSD2 | GSD3 |
| --- | --- | --- | --- |
| Deaths in first 500 days | 159–163 K | 75K | 82–83K |
| ICU capacity exceeded on | 125 days | 55–56 days | 0 days |
| Duration | 294 days | > 500 days | > 500 days |
| Susceptible after 500 days | 37–38 % | 55–56 % | 70–75% |

Compared to the benchmark scenario B2, GSD1 manages to reduce the death toll by about 70 %, merely by shielding vulnerable people (and leaving LL-interactions unrestricted). However, despite the extreme protection of vulnerable people in this scenario, ICU capacity is exceeded for 125 days and the death toll is still high (around 160 000 deaths). If the interactions involving the less vulnerable (LL) have somewhat lower contact rates (as in GSD2), we observe a lower death toll and ICU capacity is exceeded on fewer days (around 55 days). Still, there are 75 000 deaths, which is about an 85 % reduction compared to scenario B2 in which vulnerable people are not protected.

The only scenario in which ICU capacity is not exceeded is GSD3, where both groups are targeted. Compared to GSD2, this scenario does not lead to fewer deaths, though. This is due to the fact that vulnerable individuals are less restricted and, thus, those who have a higher probability to suffer a severe course of infection and to eventually pass away will incur relatively more infections. Moreover, since interactions among less vulnerable individuals are more restricted, a larger share of the population is still susceptible after 500 days. Compared to scenario USD1, where uniform social distancing is applied in a mild form and all interactions imply an effective reproduction number of about *R* = 1.5, the death toll in scenario GSD3 is reduced by about 50 000 (approx. 40 %) and ICU capacity is not exceeded, while it is exceeded on up to 11 days in scenario USD1. Hence, protection of vulnerable groups in addition to mild social distancing measures for the rest of the population can be effective to some extent. The results of scenarios GSD1 and GSD2 show, on the contrary, that protection of the vulnerable population alone is not sufficient to adequately cope with the epidemic, i.e., reduce the death toll to a bearable level and protect the health system, even if measures to protect the vulnerable population are quite extreme.

### 4.4. Tracing and Quarantine

We now use the SEI^3^Q^3^RD model to assess the effect of introducing the use of tracing apps with the aim to send individuals to quarantine who had contact to an infectious individual. Recall that we assume that only symptomatic infectious individuals are identified and that infection chains can only be broken if both, the infectious and the potentially infected individual, use the app. We investigate different levels of compliance by varying the model parameter *ψ* between 0.2 (20 % use the app, low compliance; “l”) and 0.8 (80 % use the app, high compliance; “h”). We also consider a scenario in which individuals with low vulnerability use the app more effectively. We do so against the background that (i) highly vulnerable individuals are often elderly people who might use smartphones less consistently and that (ii) the benefit of app usage among less vulnerable young people is significantly higher since it enables society to resume productive activities and, while doing so, breaking infection chains.

Tracing can be applied as the only measure or it can be combined with other strategies like uniform or group-specific social distancing. We show results for all three cases, with varying compliance patterns. The tracing scenarios we present in detail add contact tracing on top of the benchmark scenario B2 (B2-Trace), the uniform social distancing scenario USD1 (USD1-Trace) and the group-specific social distancing scenario GSD3 (GSD3-Trace). In Tables 8–10, we show results for three different patterns of compliance: both groups have high compliance, i.e., 80 % take-up rate of the app (hh), both groups have low compliance, i.e., 20 % take-up (ll), and the case in which the less vulnerable have high compliance of 80 %, while only 20% out of the vulnerable use the app effectively (hl).

**Table 8.**
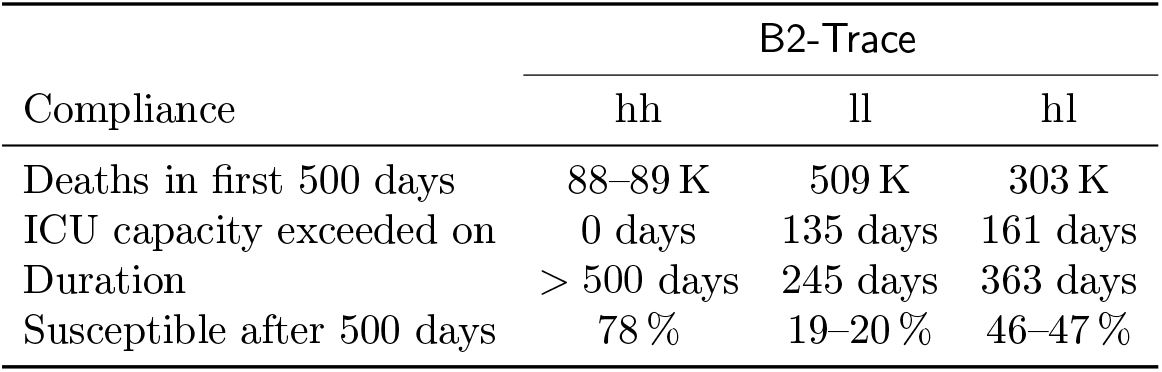
Results for the Benchmark Scenario B2 with Tracing

| Compliance | B2-Trace |  |  |
| --- | --- | --- | --- |
|  | hh | ll | hl |
| Deaths in first 500 days | 88–89 K | 509 K | 303 K |
| ICU capacity exceeded on | 0 days | 135 days | 161 days |
| Duration | > 500 days | 245 days | 363 days |
| Susceptible after 500 days | 78 % | 19–20 % | 46–47 % |

In all scenarios, tracing is effective to reduce the death toll in the first 500 days substantially if compliance is high, but not if compliance is low. In B2-Trace (only tracing without further measures), for example, with high compliance in both groups, about 450 000 deaths are avoided in the first 500 days, a reduction of about 80-85 %. Moreover, tracing is effective in keeping the number of severe cases within ICU capacity if compliance is high. There are, however, still 88 000–89 000 deaths even in this case. Recall that this scenario uses a low asymptomatic rate. The asymptomatic rate is key for tracing and if it is increased to 50 % only for the L group, then there are around 132 000 deaths in this case. With low compliance, by contrast, tracing is ineffective and barely improves over the benchmark case. At the peak, the severe cases massively outnumber ICU capacity and the death toll is high (see left panel in Figure 2). If only the group with low vulnerability is compliant, the death toll is substantially reduced compared to the same scenario without tracing. ICU capacity, however, is still exceeded for a long period of time, but less strongly compared to the case where both groups have low compliance, see the right panel in Figure 2. The fact that the epidemic progresses more slowly in this case implies that ICU capacity is exceeded for more days as compared to the case where both groups show low compliance. For the same reason it is exceeded by a smaller amount on most of these days, especially at and around the peak; see top panels of Figure 3.

**Figure 2.**
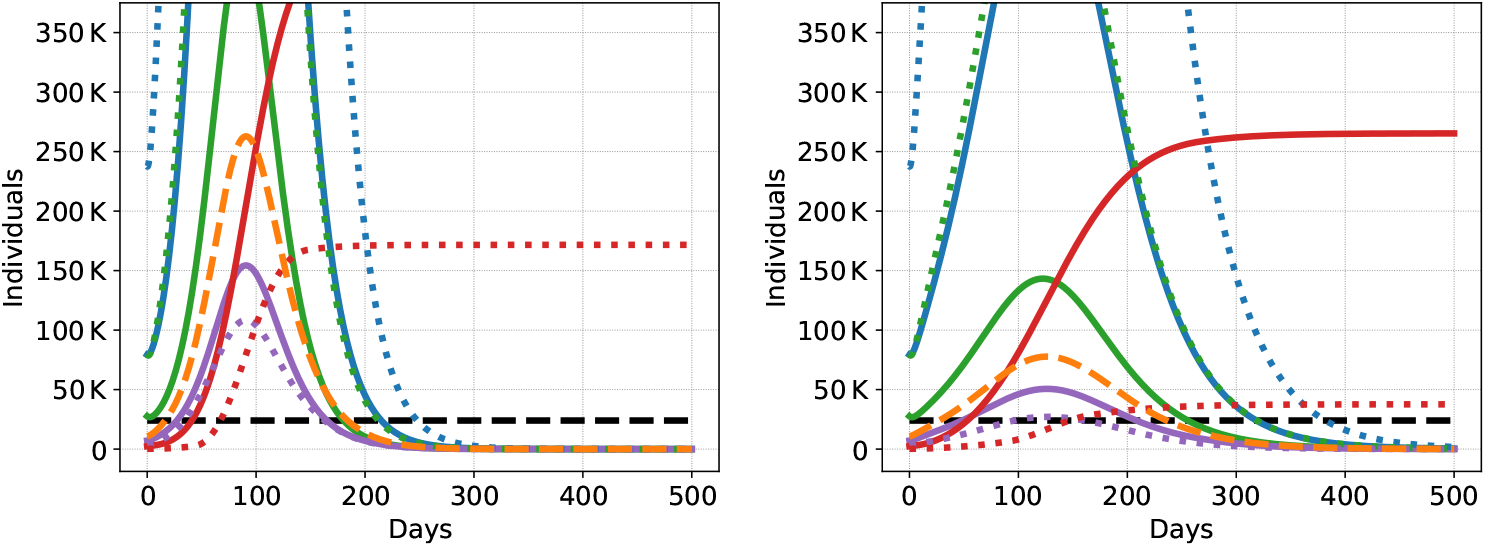
Dynamics of the COVID-19 epidemic for scenario B2-Trace with low (20%) compliance by both groups (ll, left) and with mixed (20% by group H and 80% by group L) compliance (hl, right): Symptomatic individuals (traced and not traced added together) in blue, asymptomatic in green, severe cases in purple, overall severe cases (H and L group aggregated) in orange, dead in red, and ICU capacity in black. The group with low vulnerability has dotted curves, the group with high vulnerability is plotted solidly.

**Figure 3.**
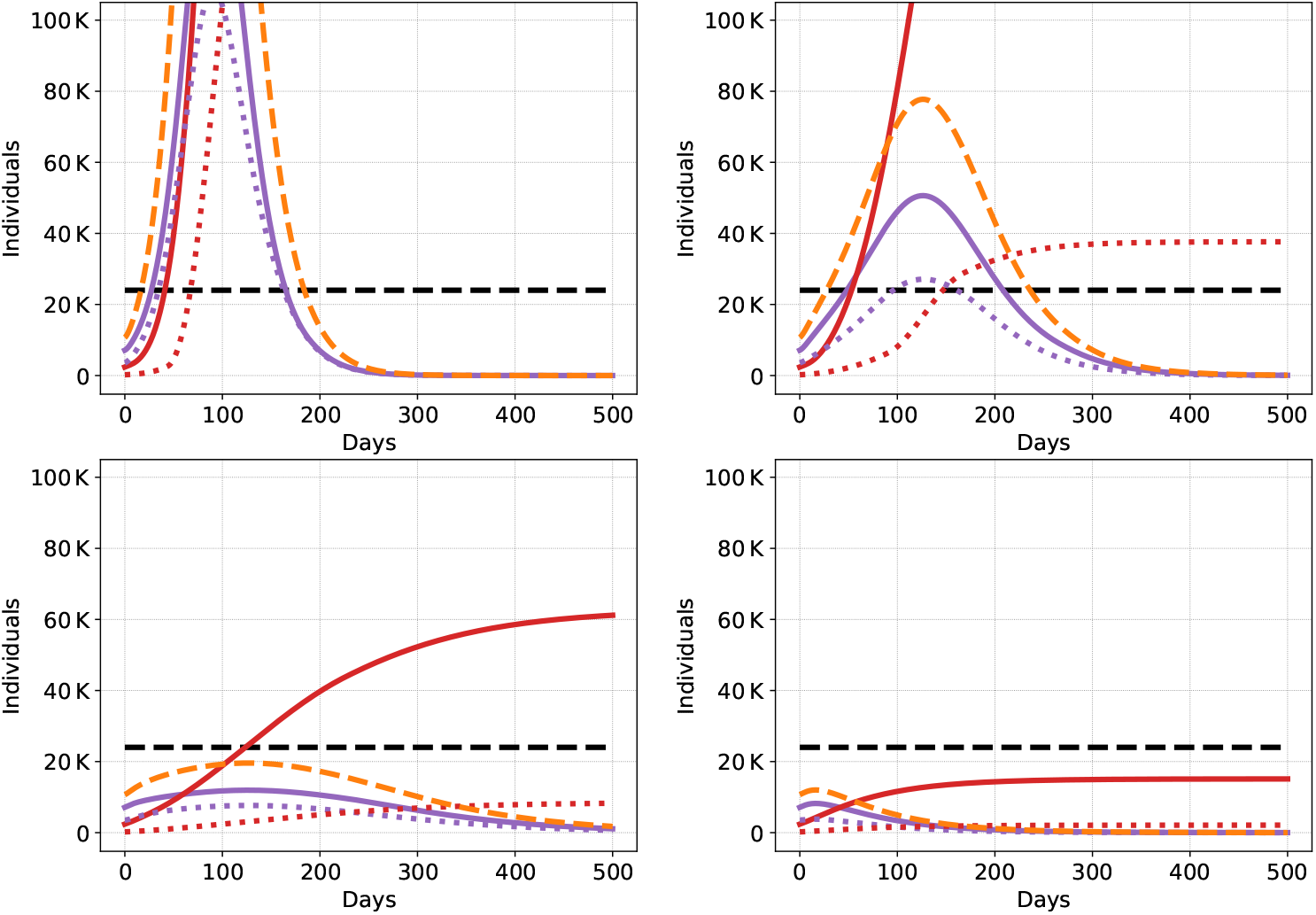
Dynamics of the epidemic (higher level of detail) for Scenario B2-Trace with low compliance (ll, top left) and mixed compliance (hl, top right) and Scenario GSD3-Trace with low compliance (ll, bottom left) and mixed compliance (hl, bottom right). Severely affected infectious individuals in purple (traced and not traced lumped together), overall severely affected infectious individuals in orange (traced and non traced of both groups, H and L), dead in red, and ICU capacity in black. The group with low vulnerability (L) has dotted curves, the group with high vulnerability (H) is plotted solidly.

If we add tracing onto a scenario with uniform or group-specific social distancing (USD1-Trace and GSD3-Trace; see Tables 9 and 10), we see similar patterns, but in a context where there is substantially less strain on the health system compared to the benchmark scenarios. The death toll is significantly reduced compared to the respective scenarios without tracing if compliance is high—but not if it is low. Interestingly, if only the group with low vulnerability has a high take-up rate, and social distancing is in place, the death toll is close to the death toll under generally high compliance if group-specific social distancing is in place. The numbers also show that even if compliance is low, tracing is able to shave off some of the peak demand for ICU beds. In scenario USD1-Trace, this means that we manage to stay within ICU capacity with tracing but not without (in the GSD3 scenario, ICU capacity is sufficient already without tracing). There are, of course, also scenarios based on group-specific social distancing in which without tracing, ICU capacity is exceeded but not without.

**Table 9.** Results for the Uniform Social Distancing Scenario USD1 with Tracing

| Compliance | USD1-Trace |  |  |
| --- | --- | --- | --- |
|  | hh | ll | hl |
| Deaths in first 500 days | 15–16 K | 79–82 K | 23–24 K |
| ICU capacity exceeded on | 0 days | 0 days | 0 days |
| Duration | > 500 days | > 500 days | > 500 days |
| Susceptible after 500 days | 98 % | 75 % | 92–93 % |

**Table 10.** Results for the Group-Specific Social Distancing Scenario GSD3 with Tracing

| Compliance | GSD3-Trace |  |  |
| --- | --- | --- | --- |
|  | hh | ll | hl |
| Deaths in first 500 days | 13–14 K | 66–69 K | 17–18 K |
| ICU capacity exceeded on | 0 days | 0 days | 0 days |
| Duration | > 500 days | > 500 days | > 500 days |
| Susceptible after 500 days | 95 % | 75–76 % | 93–94 % |

Figure 3 visualizes the difference in severe infections and death tolls in a situation where only tracing is applied without further measures (upper two figures, scenario B2-Trace) and the situation where tracing is combined with group-specific social distancing (lower two figures, scenario GSD3-Trace). The left panel shows the case of low compliance (ll), while the right panel shows the case of mixed compliance (hl). Obviously, high compliance among the individuals with low vulnerability suffices to keep the death toll extremely low in case the vulnerable group is protected (lower right panel). In case of no further measures (upper left panel), the number of severe infections are reduced, however, ICU capacity is still exceeded for a long time period and the death toll is high. This can be seen in Figure 2 (red curve), which shows the same scenario with a different scale at the ordinate.

Figure 4 highlights the crucial role of compliance when it comes to the use of tracing apps. The top-left panel shows the benchmark case B2-Trace. It illustrates that—if compliance is high—the death toll can be significantly reduced. The figure also illustrates a very steep gradient. As compliance decreases, the death toll increases very quickly. For a compliance rate of 70 %, the number of deaths exceeds 200 000 (a 1700 % increase over full compliance) and as compliance drops to 40 %, the number of deaths shoots up to exceed 500 000. This suggests that tracing should be used in a careful way and combined with a tailored strategy to open up sectors where tracing can be made mandatory—if one wants to rely on its effectiveness. The bottom-left panel shows the same effects in scenario GSD3-Trace. As social distancing measures are in place in this scenario, the death toll is smaller. Note also that—since vulnerable groups are protected in this scenario—the gradient is less steep: 70 % compliance in this scenario implies a 70 % increase in deaths compared to full compliance, as opposed to the 1700 % increase observed in B2-Trace.

**Figure 4.**
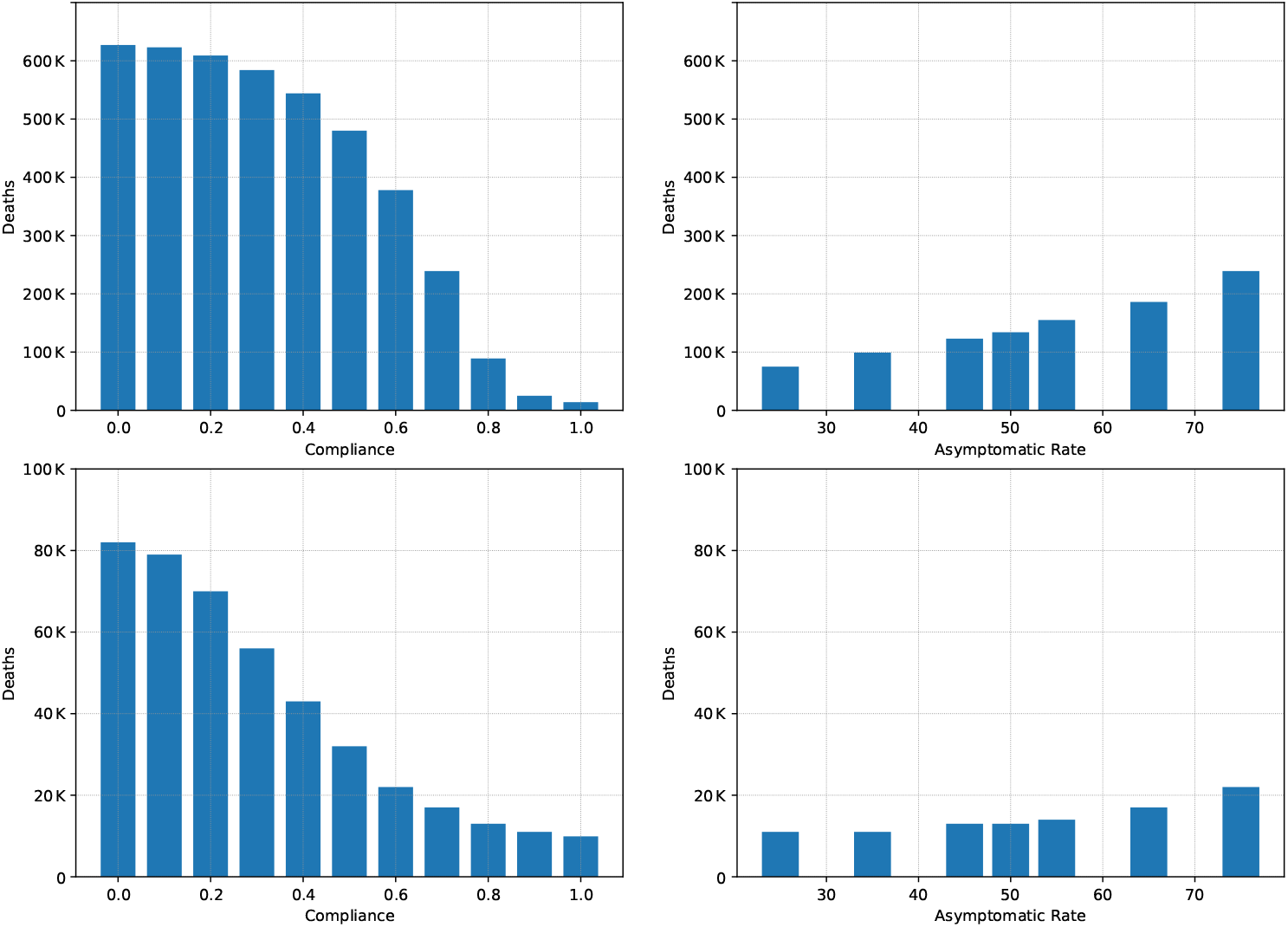
Role of Compliance and the Asymptomatic Rate for Tracing. Panel (top-left): Changing Compliance in B2. Panel (top-right): Changing the Asymptomatic Rate in B2 with high (80%) compliance. (bottom-left): Changing Compliance in GSD3. Panel (bottom-right): Changing the Asymptomatic Rate in GSD3 with high (80%) compliance.

Another crucial parameter for the success of tracing is the asymptomatic rate. As asymptomatic individuals do not notice that they are ill, they and their contacts cannot be removed from the infectious population via contact tracing. The top-right panel in Figure 4 shows the effect of increasing the asymptomatic rate in Scenario B2 with 80 % compliance, starting from 25 %. Clearly, the higher the asymptomatic rate the less effective tracing is in reducing new infections and ultimately deaths. The bottom-right panel shows a similar effect for GSD3. It is noticeable, though, that the gradient here is much less steep, i.e., contact tracing with a high compliance rate is still effective if the asymptomatic rate is higher.

Last, Appendix Figures 5 and 6 show the same curves on the same scale as Figure 1 for the scenarios discussed in Figures 2 and 3.

**Figure 5.**
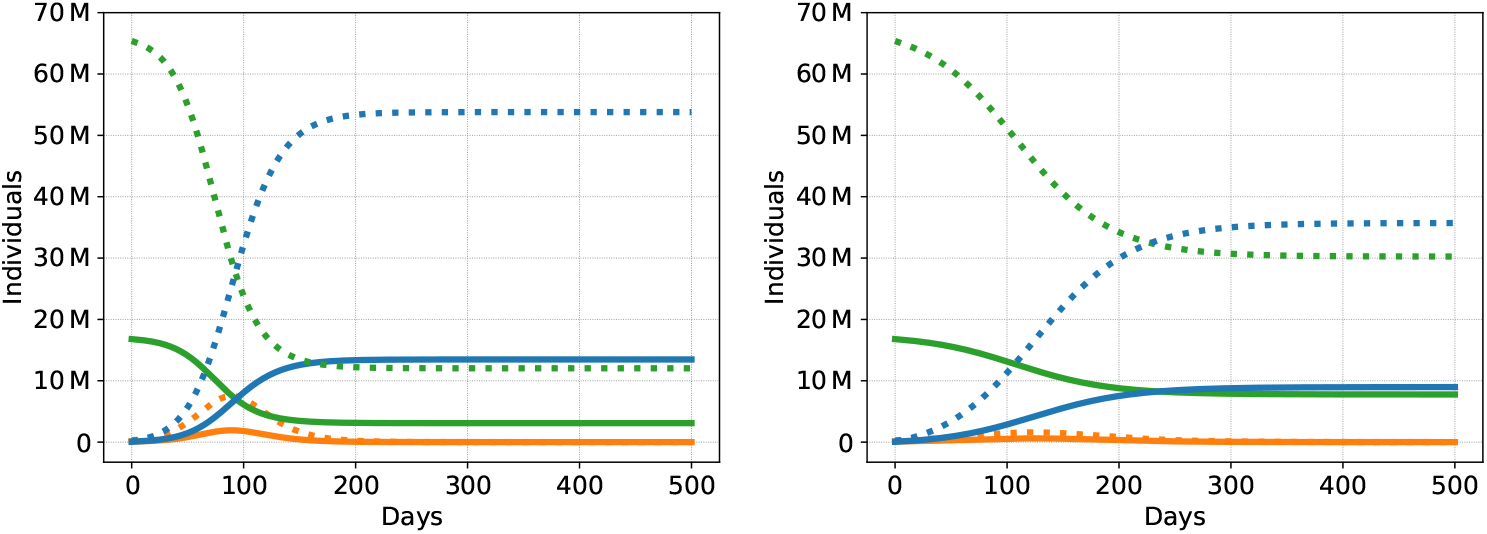
A larger view of dynamics of the COVID-19 epidemic for scenario B2-Trace with low (20%) compliance by both groups (ll, left) and with mixed (20% by group H and 80% by group L) compliance (hl, right): Symptomatic individuals (traced and not traced added together) in blue, asymptomatic in green, severe cases in purple, overall severe cases (H and L group aggregated) in orange, dead in red, and ICU capacity in black. The group with low vulnerability has dotted curves, the group with high vulnerability is plotted solidly.

**Figure 6.**
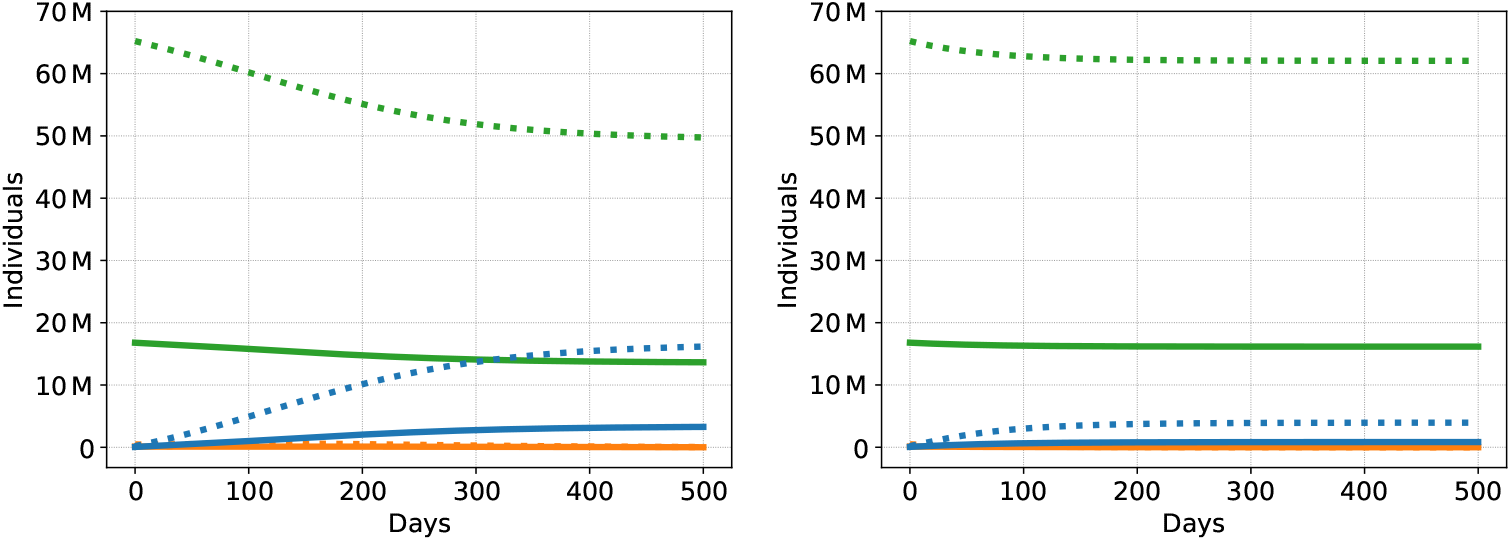
A larger view of dynamics of the COVID-19 epidemic for scenario GSD3-Trace with low (20%) compliance by both groups (ll, left) and with mixed (20% by group H and 80% by group L) compliance (hl, right): Symptomatic individuals (traced and not traced added together) in blue, asymptomatic in green, severe cases in purple, overall severe cases (H and L group aggregated) in orange, dead in red, and ICU capacity in black. The group with low vulnerability has dotted curves, the group with high vulnerability is plotted solidly.

## 5. Conclusion

In this paper, we propose an extension of the epidemiological SEIR model to enable the analysis of several measures of epidemic control that are currently discussed. We introduce different groups into the SEIR model that may differ both in their underlying parameters as well as in their behavioral response to public health interventions. In particular, we allow the infectiousness parameters to differ within and across groups. Moreover, groups may have different asymptomatic, hospitalization, and lethality rates. Within this framework, we are able to analyze social distancing measures that differentiate across groups, specific protection of groups with high lethality rates, as well as the detailed effects of “trace & isolate” strategies with different levels of take-up of the tracing apps needed. We moreover shed light on the attractiveness of combining these measures.

Our results highlight sharp trade-offs between different goals of epidemic control and also illustrate how tailored social distancing and protection measures could be effectively combined with a “trace & isolate” approach to keep severe infections within the capacity of the health system and the death toll low. However, our analysis also clearly illustrates the well known downside of rigorous epidemic control: The duration of the epidemic is extended far into the future, making it necessary to continue rigorous control measures for possibly very long time-spans until a vaccine becomes widely available. On the positive side, we illustrate that mild social distancing in combination with a “trace & isolate” approach could be a promising way to control the epidemic in the medium run. This is, however, only true if take-up rates for the apps are high enough within those groups in which social distancing restrictions are looser. This is an unlikely scenario in everyday life, however, it could well create opportunities to open businesses under the condition that tracing is made mandatory. This could definitely apply to industrial production and the services industry, and may also be a temporary regime in education. From a practical perspective, the speed of infection control will also be crucial for effectiveness. In our model we assume that infection chains are broken whenever the infectious and the infected individual use an app. This, however, is only true if the health system operates fast enough to isolate the infected individual within the incubation period. Let us finally note that all our results stem from a case study that necessarily builds on a fragile data base. As we learn more from randomized controlled trials and large-scale observational studies, this situation will change. The main contribution of our paper, the extension of the SEIR model to SEI^3^Q^3^RD, can easily be applied to new and more reliable data sets. Our model and results also provide guidance to which parameters are key to be identified reliably to analyze the effectiveness of epidemic control approaches in more detail.

## Data Availability

The data used in the paper is also explicitly described in the paper.

## Acknowledgments

The authors thank the Deutsche Forschungsgemeinschaft for their support within project A05 (Schmidt) and B08 (Grimm, Schmidt) in the “Sonderforschungsbereich/Transregio 154 Mathematical Modelling, Simulation and Optimization using the Example of Gas Networks”, as well as for their support within the project “The role of social identity for learning in networks” (Grimm, Mengel). We also acknowledge funding by the Bavarian State Government for the research initiative Energie Campus Nürnberg (Grimm). Finally, we thank Benjamin Buschmann, Julia Markovsek and Karolína Predotová for valuable research assistance.

## Author Contribution Statements

Veronika Grimm, Friederike Mengel, and Martin Schmidt all contributed to the writing of the main manuscript text as well as to the production of the figures and the tables.

## Additional Information

The authors declare no competing interests.

## 6. Additional Figures

## Notes

### Competing Interest Statement

The authors have declared no competing interest.

### Funding Statement

No external funding was explicitly received for any aspect of the submitted work.

### Summary of Updates

Some data has been updated and thus, some numerical results quantitatively changed as well. However, the qualitative results remain the same. Further improvements in the presentation of the results have been incorporated.

